# Estimating the period prevalence of SARS-CoV-2 infection during the Omicron (BA.1) surge in New York City (NYC), January 1-March 16, 2022

**DOI:** 10.1101/2022.04.23.22274214

**Authors:** Saba A Qasmieh, McKaylee M Robertson, Chloe A Teasdale, Sarah G Kulkarni, Denis Nash

**Author notes:** Correspondence to: Saba Qasmieh, MPH, Institute of Implementation Science in Population Health, City University of New York, New York, NY.

## Abstract

In a population-based survey of NYC adults, we assessed positive SARS-CoV-2 tests (including via exclusive at-home testing) and possible cases among untested respondents. An estimated 27.4% (95%CI: 22.8%-32.0%) or 1.8 million adults (95%CI: 1.6-2.1 million) had SARS-CoV-2 infection. SARS-CoV-2 prevalence was high among groups that are more vulnerable to severe SARS-CoV-2 and death, including unvaccinated persons (21.7%, 95%CI 9.6%-33.8%) and those aged 65+ (17.8%, 95%CI 10.2-25.4%). Population-based representative surveys are an important adjunct surveillance tool to standard testing-based SARS-CoV-2 surveillance.

## Introduction

Routine case-based surveillance data on individuals who test or present to care for SARS-CoV-2 underestimate the true burden of SARS-CoV-2 infection in the general population due to 1) undiagnosed/untested cases^1^; and 2) the exclusive use of at-home rapid tests which are not reflected in routine case surveillance in the U.S.^2^ The degree of underestimation could be differential by geographic and sociodemographic factors, and vary over time.^3,4^ Concerns raised about the potential for biased interpretation of SARS-CoV-2 case counts, case rates, and test positivity rates recently led the Centers of Disease Control and Prevention (CDC) to update their guidelines for community COVID-19 metrics to inform community prevention measures, shifting emphasis to hospital admissions and deaths.^2^ While important, elevated hospital admissions and deaths resulting from a surge lag behind increases in community transmission, resulting in missed opportunities for earlier mitigation of a surge.

Despite these national shifts in emphasis, the number of new cases and the test positivity rates among SARS-CoV-2 testers are still relied on by local governments, citizens, and the news media to infer levels of SARS-CoV-2 community transmission and trigger action. This could be increasingly problematic, especially early in surges, due to time-varying factors such as decreased test-seeking behaviors and increased access to and availability of at-home testing, which complicate interpretation of these metrics.^5^

This study aimed to assess the extent to which routine case surveillance underestimated the burden of SARS-CoV-2 infections during the recent Omicron surge.

## Methods

We conducted a cross-sectional survey March 14-16, 2022, of 1,030 adult New York City (NYC) residents. Respondents were asked about SARS-CoV-2 testing and related outcomes *since January 1, 2022*, which represents the second half of the Omicron BA.1 surge in NYC. Survey weights were applied to generate estimates for non-institutionalized NYC residents aged 18+. Additional details on the survey design are included in the *Statistical Appendix*. The study protocol was approved by the Institutional Review Board at the City University of New York (CUNY).

### Period Prevalence Estimation

The survey questionnaire ascertained the types and results of viral diagnostic tests taken between January 1, 2022, and survey date (March 14-16, 2022), including PCR, rapid antigen and/or at-home rapid tests. The survey also captured information on COVID-19 symptoms among respondents during this time period, as well as known close contacts with a confirmed or probable case of SARS-CoV-2 infection. COVID-19 symptoms included any of the following: fever of 100° F or greater, cough, runny nose and/or nasal congestion, shortness of breath, sore throat, fatigue, muscle/body aches, headaches, loss of smell/taste, nausea, vomiting and/or diarrhea.^6^

We estimated the number and proportion of respondents who likely had SARS-CoV-2 infection during the study period based on the following mutually exclusive, hierarchical case classification: 1) Confirmed case: self-report of one or more positive tests with a health care or testing provider; or 2) Probable case: self-report of a positive test result exclusively on at-home rapid tests (i.e. those that were not followed up with confirmatory diagnostic testing with a provider); or 3) Possible case: self-report of COVID-like symptoms <u>and</u> a known epidemiologic link (close contact) to one or more laboratory confirmed or probable (symptomatic) SARS-CoV-2 case(s)^6^ in a respondent who reported never testing or only testing negative during the study period. Categories 1 and 2 of our case definition would likely capture some, but not all, of the estimated 20-30% of individuals whose SARS-CoV-2 infection may remain asymptomatic throughout their infection.^7^

### Statistical Analysis

We described the testing status and estimated period prevalence of SARS-CoV-2 by socio-demographic characteristics, geography, and vaccination status. Pearson’s chi-squared test of independence was performed to assess group differences between testers and non-testers. Analyses were performed using SAS version 9.4.

## Results

We estimate that 27.4% (95% CI22.8%-32.0%) of approximately 6.6 million adult New Yorkers may have had SARS-CoV-2 infection during January 1-March 16, 2022, corresponding to about 1.8 million adults (95%CI 1.6-2.1 million). The estimate of 27.4% includes: 1) 14.1% (95%CI 10.4%-17.8%) who were positive based on one or more tests with a health care or testing provider; 2) 5.2% (95%CI 3.1%-7.3%) who were positive exclusively based on one or more at-home rapid tests; and 3) 8.1% (95%CI 5.4%-10.9%) who met the definition for possible SARS-CoV-2 infection. The test positivity rate among those who tested with a healthcare or testing provider was 41.3% (95%CI 33.2% - 49.4%).

The weighted characteristics of survey participants and period prevalence estimates are shown in Table 1. SARS-CoV-2 period prevalence during this was high among all groups, but varied substantially by sociodemographic factors and geography. Importantly, SARS-CoV-2 prevalence was high among groups that are more vulnerable to severe SARS-CoV-2 and death, including unvaccinated persons (21.7%, 95%CI 9.6%-33.8%) and those aged 65+ (17.8%, 95%CI 10.2-25.4%). Individuals who tested at all were more likely to be 18-34 years, Hispanic, and have higher education levels and combined household income >$65K compared with those who did not test at all.

**Table 1.**
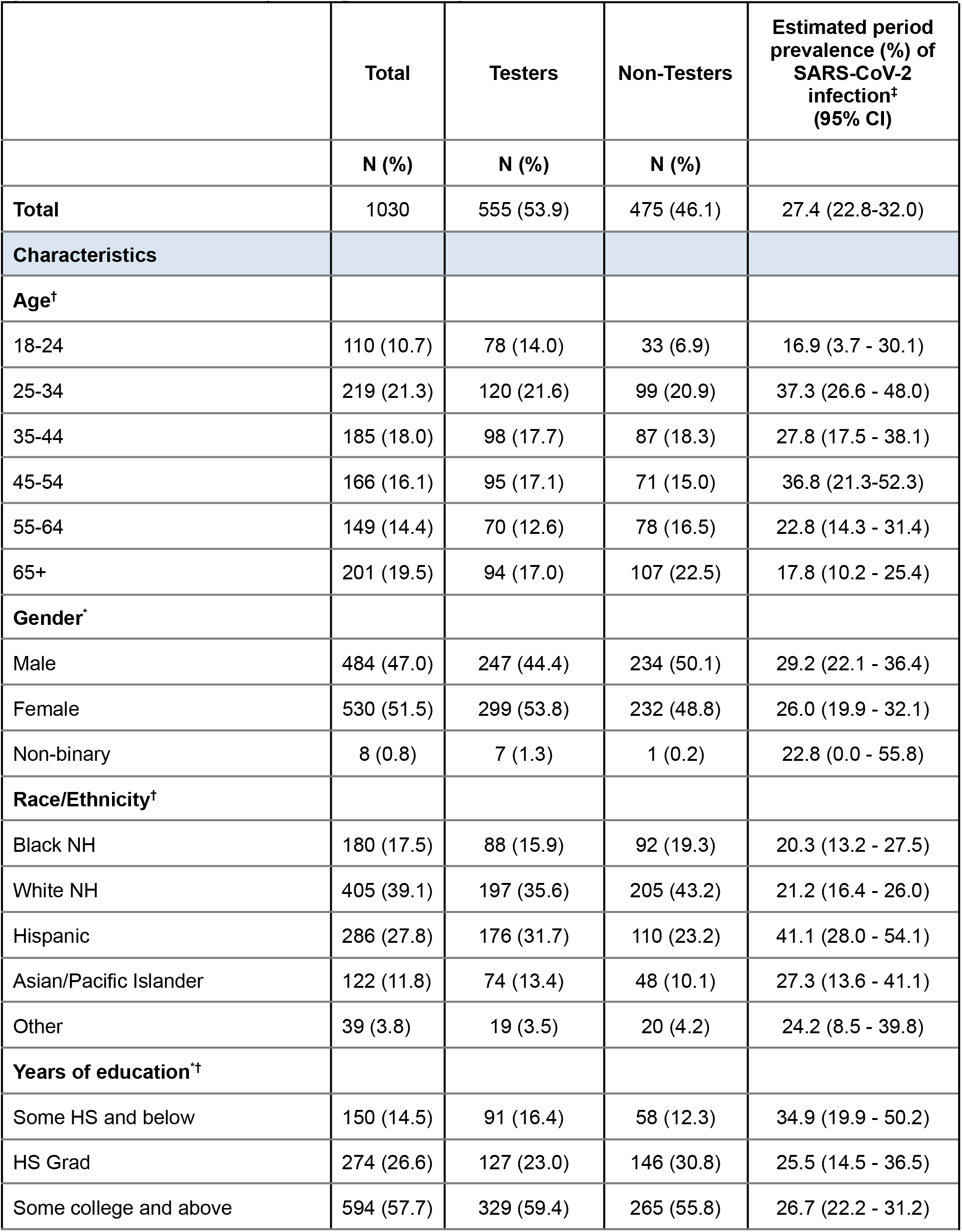
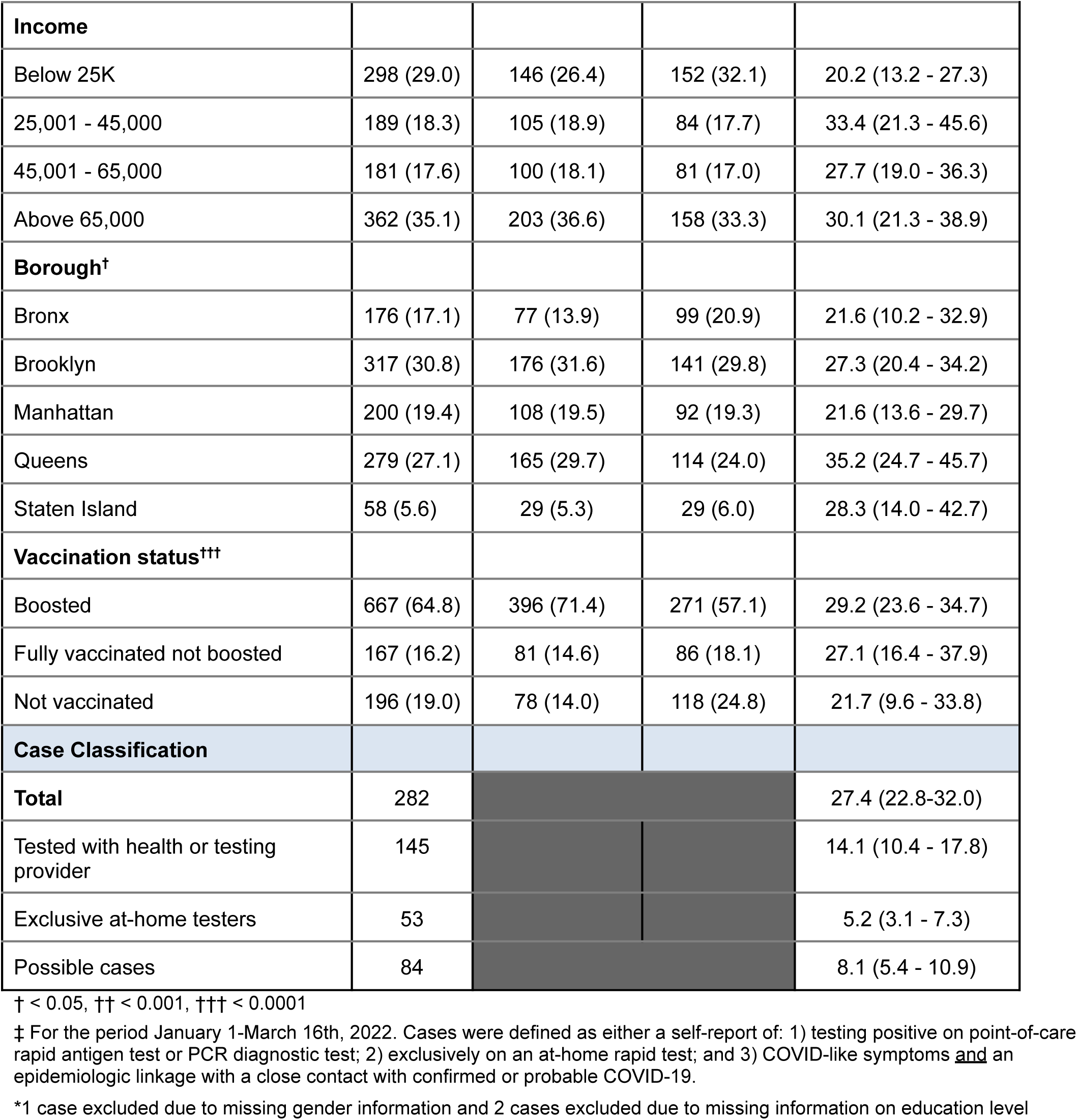
Select socio-demographic characteristics for survey respondents by testing status and period prevalence of SARS-CoV-2, January-March 16^th^, 2022.

|  | Total | Testers | Non-Testers | Estimated period prevalence (%) of SARS-CoV-2 infection<sup>†</sup> (95% CI) |
| --- | --- | --- | --- | --- |
|  | N (%) | N (%) | N (%) |  |
| **Total** | 1030 | 555 (53.9) | 475 (46.1) | 27.4 (22.8-32.0) |
| **Characteristics** |  |  |  |  |
| **Age<sup>†</sup>** |  |  |  |  |
| 18-24 | 110 (10.7) | 78 (14.0) | 33 (6.9) | 16.9 (3.7 - 30.1) |
| 25-34 | 219 (21.3) | 120 (21.6) | 99 (20.9) | 37.3 (26.6 - 48.0) |
| 35-44 | 185 (18.0) | 98 (17.7) | 87 (18.3) | 27.8 (17.5 - 38.1) |
| 45-54 | 166 (16.1) | 95 (17.1) | 71 (15.0) | 36.8 (21.3-52.3) |
| 55-64 | 149 (14.4) | 70 (12.6) | 78 (16.5) | 22.8 (14.3 - 31.4) |
| 65+ | 201 (19.5) | 94 (17.0) | 107 (22.5) | 17.8 (10.2 - 25.4) |
| **Gender<sup>\*</sup>** |  |  |  |  |
| Male | 484 (47.0) | 247 (44.4) | 234 (50.1) | 29.2 (22.1 - 36.4) |
| Female | 530 (51.5) | 299 (53.8) | 232 (48.8) | 26.0 (19.9 - 32.1) |
| Non-binary | 8 (0.8) | 7 (1.3) | 1 (0.2) | 22.8 (0.0 - 55.8) |
| **Race/Ethnicity<sup>†</sup>** |  |  |  |  |
| Black NH | 180 (17.5) | 88 (15.9) | 92 (19.3) | 20.3 (13.2 - 27.5) |
| White NH | 405 (39.1) | 197 (35.6) | 205 (43.2) | 21.2 (16.4 - 26.0) |
| Hispanic | 286 (27.8) | 176 (31.7) | 110 (23.2) | 41.1 (28.0 - 54.1) |
| Asian/Pacific Islander | 122 (11.8) | 74 (13.4) | 48 (10.1) | 27.3 (13.6 - 41.1) |
| Other | 39 (3.8) | 19 (3.5) | 20 (4.2) | 24.2 (8.5 - 39.8) |
| **Years of education<sup>††</sup>** |  |  |  |  |
| Some HS and below | 150 (14.5) | 91 (16.4) | 58 (12.3) | 34.9 (19.9 - 50.2) |
| HS Grad | 274 (26.6) | 127 (23.0) | 146 (30.8) | 25.5 (14.5 - 36.5) |
| Some college and above | 594 (57.7) | 329 (59.4) | 265 (55.8) | 26.7 (22.2 - 31.2) |

| **Income** |  |  |  |  |
| --- | --- | --- | --- | --- |
| Below 25K | 298 (29.0) | 146 (26.4) | 152 (32.1) | 20.2 (13.2 - 27.3) |
| 25,001 - 45,000 | 189 (18.3) | 105 (18.9) | 84 (17.7) | 33.4 (21.3 - 45.6) |
| 45,001 - 65,000 | 181 (17.6) | 100 (18.1) | 81 (17.0) | 27.7 (19.0 - 36.3) |
| Above 65,000 | 362 (35.1) | 203 (36.6) | 158 (33.3) | 30.1 (21.3 - 38.9) |
| **Borough<sup>†</sup>** |  |  |  |  |
| Bronx | 176 (17.1) | 77 (13.9) | 99 (20.9) | 21.6 (10.2 - 32.9) |
| Brooklyn | 317 (30.8) | 176 (31.6) | 141 (29.8) | 27.3 (20.4 - 34.2) |
| Manhattan | 200 (19.4) | 108 (19.5) | 92 (19.3) | 21.6 (13.6 - 29.7) |
| Queens | 279 (27.1) | 165 (29.7) | 114 (24.0) | 35.2 (24.7 - 45.7) |
| Staten Island | 58 (5.6) | 29 (5.3) | 29 (6.0) | 28.3 (14.0 - 42.7) |
| **Vaccination status<sup>†††</sup>** |  |  |  |  |
| Boosted | 667 (64.8) | 396 (71.4) | 271 (57.1) | 29.2 (23.6 - 34.7) |
| Fully vaccinated not boosted | 167 (16.2) | 81 (14.6) | 86 (18.1) | 27.1 (16.4 - 37.9) |
| Not vaccinated | 196 (19.0) | 78 (14.0) | 118 (24.8) | 21.7 (9.6 - 33.8) |
| **Case Classification** |  |  |  |  |
| **Total** | 282 |  |  | 27.4 (22.8-32.0) |
| Tested with health or testing provider | 145 |  |  | 14.1 (10.4 - 17.8) |
| Exclusive at-home testers | 53 |  |  | 5.2 (3.1 - 7.3) |
| Possible cases | 84 |  |  | 8.1 (5.4 - 10.9) |

† < 0.05, †† < 0.001, ††† < 0.0001
‡ For the period January 1-March 16th, 2022. Cases were defined as either a self-report of: 1) testing positive on point-of-care rapid antigen test or PCR diagnostic test; 2) exclusively on an at-home rapid test; and 3) COVID-like symptoms and an epidemiologic linkage with a close contact with confirmed or probable COVID-19.
\*1 case excluded due to missing gender information and 2 cases excluded due to missing information on education level

## Discussion

Our study found a high prevalence of SARS-CoV-2 among adult New Yorkers during the second half of the city’s Omicron BA.1 surge in January-mid March 2022 (27.4% or about 1.8 million people), including among groups who are at higher risk for a severe outcome of SARS-CoV-2 infection. We also found that the characteristics of adult New Yorkers testing with a health care or testing provider differ substantially from those who do not test, highlighting the challenges of using surveillance data solely based on testing to gain insights into the burden and epidemiology of SARS-CoV-2 community transmission.

During the study period, routine case surveillance data from the NYC DOHMH reported 552,084 NYC residents of all ages (∼6.7% of the NYC population of 8.3M) who tested positive for SARS-CoV-2 with a health care or testing provider by PCR or point of care rapid test.^8^ The 7-day average test percent positivity during the study period ranged from 34.8% on January 1st to 1.6% on March 16th with 11.5% percent positivity among total testers during the entire period.^8^ When compared with our estimate of 1.8 million adults infected during the same time period, our findings point to the extent to which official case counts underestimated the SARS-CoV-2 burden during the surge. This ‘hidden prevalence’ is due to both non-testing, exclusive at-home rapid testing, and testing too soon after exposure/symptom onset with either a point of care and at-home rapid tests.^1,9^

The recent CDC metrics may be inadequate for informing timely public health countermeasures, including protecting and reaching those who are most at risk of a severe outcome. While wastewater surveillance^10^ is an important tool for early detection of a surge, surveillance methods like routinely and strategically deployed surveys enables an assessment of the prevalence of infection as well as an assessment of the characteristics of populations, including vulnerable populations, that are infected, thereby providing critical epidemiologic intelligence between wastewater signals and possible future spikes in hospitalizations.

While our study suggests a viable and simple approach to gather important and timely information about the prevalence of SARS-CoV-2 infections among adults in NYC, it also has limitations. First, we measured testing outcomes and symptoms via self-report over a long recall period, which is subject to recall bias. More frequent surveys with shorter recall periods (e.g., 7-14 days), could improve the accuracy of estimates. Our prevalence estimates also included possible SARS-CoV-2 cases based on self-reported symptoms who had a known contact with a confirmed/probable case, which, even though both prevalence of exposures and attack rates were very high during the BA.1 Omicron surge^11^, could lead to an overestimate of prevalence. Conversely, our estimates may not have captured some SARS-CoV-2 cases that are asymptomatic for their entire infection, resulting in an underestimate (e.g., by 10-30%).^7^ In addition, our survey excludes children and adolescents <18, those who died (about 4,426 NYC residents) during the study period. The small sample size limits the precision of some estimates across respondent characteristics. Part of our sample (32%) was derived from online panel data, as opposed to a population-based sampling frame (see Statistical Appendix), which could introduce bias. Finally, those who chose to participate in the survey could be more or less likely to have had COVID-19 recently (participation bias).

Population-based representative surveys are an important adjunct surveillance tool to standard testing-based SARS-CoV-2 surveillance. At this stage of the pandemic, the application of low-cost and low-resource intensive surveys may have a large impact on the efforts of governments and individuals to control and prevent community spread of SARS-CoV-2, as well as secondary prevention of severe SARS-CoV-2 outcomes. Future surveys should capture additional detail on vulnerability to a severe COVID-19 outcome among those with SARS-CoV-2 infection.

## Data Availability

All data produced in the present study are available upon reasonable request to the authors.

## Acknowledgements

The authors wish to acknowledge the survey participants and Consensus Strategies for completing survey sampling and data collection.

## Statistical Appendix

### Sampling Frame

Sampling frames of NYC residents, one of 2,185,659 NYC residents with mobile numbers and one of an additional 1,532,518 with landlines, were used. From these sampling frames, two stratified proportionate random samples were drawn, one from the mobile number frame and one from the landline frame. These two samples comprised 68% of the larger study sample. The remaining 32% of the study sample came from an opt-in Online Panel provided by Consensus Strategies was used to supplement underrepresented populations in the main study sample. The final sample included n=1,030 and was weighted to the adult NYC population (details below).

### Multi-mode data collection design

Short message service (SMS) aka text messages were sent using SMS platform. The respondents were sent a personalized first name text message which included a link to the survey and an opt-out option. The respondents had the option to reply to the SMS text with any queries. Data was verified by IP address and scrubbed against the original survey sample.

Interactive voice response (IVR) aka robo-poll messages were sent to landlines using a voice recorded IVR platform. The respondents were able to answer the survey questions using the touch tone keypad on their phones.

The opt-in online panel was created by Consensus Strategies and participants were paid an incentive to complete the surveys of up to $2. Respondents were verified by payment information.

## Mode of data collection

|  |  | Frequency | Percent | Valid Percent | Cumulative Percent |
| --- | --- | --- | --- | --- | --- |
| Valid | Landline | 409 | 39.7 | 39.7 | 39.7 |
|  | Online | 334 | 32.4 | 32.4 | 72.1 |
|  | Mobile | 287 | 27.9 | 27.9 | 100.0 |
|  | Total | 1030 | 100.0 | 100.0 |  |

Data was collected March 14-16, 2022.

### Survey weighting

To account for differences in the distribution by groups the following demographic weights were developed based on the American Community Survey 1 year estimates and US Census data. These included respondent self-identified sex, educational attainment, age, ethnicity/race and region. The samples (landline, online, mobile) were normalized at the borough level based on sex, age, gender, education, race and sample size then combined and weighted back based on the proportion of the borough to the overall population and the other demographics. All variables were weighted at the same time for each borough and then the five boroughs were recombined to create the overall sample for NYC. Weights were created with IBM’s SPSS software. The inference population is 6.6 million adult NYC residents.

## Survey questionnaire

### Survey on recent COVID exposure, COVID infection, and testing behaviors in New York City

*<u>The following questions will ask about demographic characteristics</u>*

1. Which zip code do you reside in?
2. What is your age?
3. How do you currently identify your gender? Do you identify as …
  a. Male
  b. Female
  c. Gender non-binary
  d. Don’t know/not sure
4. Are you Latino/a, or of Hispanic or Spanish origin?
  a. Yes
  b. No
  c. Don’t know/not sure
5. Which one of the following would you use to describe yourself ?
  a. White
  b. Black or Black American
  c. Asian, Native Hawaiian or Other Pacific Islander
  d. American Indian, Native, First Nations, Indigenous Peoples of the Americas, or Alaska Native
  e. More than one race
  f. Don’t know/not sure
6. What is the highest grade or year of school you completed?
  a. Never attended school or only attended kindergarten
  b. Grades 1 through 8 (Elementary)
  c. Grades 9 through 11 (Some high school)
  d. Grade 12 or GED (High school graduate)
  e. College 1 year to 3 years (Some college or technical school, associate degree)
  f. College 4 years or more (College graduate)
  g. Don’t know/not sure
7. How many members of your household, including yourself, are 18 years of age or older?
8. How many children 17 years old or younger usually live or stay with you?
9. Are you currently employed for wages or salary?
  a. Yes
  b. No
  c. Don’t know/not sure
  d. Refused
10. What is your household’s annual income?
  a. Below $15,000
  b. Between $15,001 - $25,000
  c. Between $25,001 - $35,000
  d. Between $35,001 - $45,000
  e. Between $45,001 - $55,000
  f. Between $55,001 - $65,000
  g. Between $65,001 - $75,000
  h. Above $75,0000 *<u>The following questions will ask about your COVID exposure</u>* ***<u>since January 1</u>***^***<u>st</u>***^, ***<u>2022</u>***
11. Since January 1, 2022, did you experience any COVID-like symptoms (e.g., 100 degrees fever, chills, cough, sore throat, fatigue, headache, shortness of breath, congestion or runny nose, muscle aches, loss of smell or taste, nausea, or diarrhea)?
  a. Yes
  b. No
  c. Don’t know/not sure
12. Since January 1, 2022, not including yourself, did anyone in your household either experience COVID-like symptoms or has tested positive for COVID-19?
  a. Yes
  b. No
  c. Don’t know/not sure
13. Since January 1, 2022, were you exposed to any other person <u>outside of your household</u> who either had COVID-like symptoms or tested positive for COVID-19?
  a. Yes
  b. No
  c. Don’t know/not sure
14. Have you been fully vaccinated against COVID-19 with a vaccine that has received FDA approval or emergency use authorization?
  a. Yes
  b. No
  c. Don’t know/not sure
15. If you have been fully vaccinated, have you received a coronavirus booster?
  a. Yes, before January 1, 2022
  b. Yes, after January 1, 2022
  c. No
  d. Don’t know/not sure
16. Do you have any kind of health care coverage, including health insurance, prepaid plans such as HMOs, or government plans such as Medicare, or Indian Health Service?
  a. Yes
  b. No
  c. Don’t know/not sure
17. Since January 1, 2022, do you <u>think</u> you had a COVID-19 infection?
  a. Yes
  b. No
  c. Don’t know/not sure *<u>The following questions will ask about COVID-19 testing</u>* ***<u>since January 1</u>***^***<u>st</u>***^, ***<u>2022</u>***
18. Since January 1, 2022, have you tested positive for COVID-19 on an at-home rapid test (a rapid at-home test allows you to collect your own sample and get results within minutes at home)?
  1. Yes
  2. No
  c. Don’t know/not sure
19. Since January 1, 2022, have you tested negative for COVID-19 on an at-home rapid test (a rapid at-home test allows you to collect your own sample and get results within minutes at home)?
  a. Yes
  b. No
  c. Don’t know/not sure
20. Since January 1, 2022, have you tested positive for COVID-19 on a rapid antigen or a PCR test from a healthcare or testing provider?
  a. Yes
  b. No
  c. Don’t know/not sure
21. Since January 1, 2022, have you tested negative for COVID-19 on a rapid antigen or a PCR test from a healthcare or testing provider?
  a. Yes
  b. No
  c. Don’t know/not sure
22. *If you tested positive on any viral test (at home-self test, or rapid or PCR at a healthcare or testing provider) since January 1st, did you test positive in the last 7 days?*
  a. Yes
  b. No
  c. Don’t know/not sure
23. Since January 1, 2022, did you have any difficulty in getting a viral COVID-19 test (*PCR or rapid at a healthcare or testing provider*) for <u>yourself</u>?
  a. I had no difficulty in getting a viral COVID-19 test
  b. I had some difficulty in getting a viral COVID-19 test
  c. I had a lot of difficulty in getting a viral COVID-19 test
  d. I did not seek a viral COVID-19 test
  e. Don’t know/not sure <u>*The following questions will ask about the locations where you may have tested for COVID-19*</u> ***<u>since January 1</u>***^***<u>st</u>***^, ***<u>2022</u>***
24. Since January 1, 2022, have you tested for COVID-19 at a hospital or a physician’s office?
  a. Yes, and tested positive
  b. Yes, but tested negative
  c. No
  d. Don’t know/not sure
  e. Refused
25. Since January 1, 2022, have you tested for COVID-19 at a CityMD clinic?
  a. Yes, and tested positive
  b. Yes, but tested negative
  c. No
  d. Don’t know/not sure
  e. Refused
26. Since January 1, 2022, have you tested for COVID-19 at an urgent care clinic that is not CityMD?
  a. Yes, and tested positive
  b. Yes, but tested negative
  c. No
  d. Don’t know/not sure
  e. Refused
27. Since January 1, 2022, have you tested for COVID-19 at a mobile testing site?
  a. Yes, and tested positive
  b. Yes, but tested negative
  c. No
  d. Don’t know/not sure
  e. Refused
28. Since January 1, 2022, have you tested for COVID-19 at a pharmacy?
  a. Yes, and tested positive
  b. Yes, but tested negative
  c. No
  d. Don’t know/not sure
  e. Refused

